# Single cell transcriptomic analysis of diffuse large B cells in cerebrospinal fluid of central nervous system lymphoma

**DOI:** 10.1101/2020.04.18.20065805

**Authors:** Haoyu Ruan, Zhe Wang, Yue Zhai, Ying Xu, Linyu Pi, Jihong Zheng, Yihang Zhou, Cong Zhang, RuoFan Huang, Kun Chen, Xiangyu Li, Weizhe Ma, Zhiyuan Wu, Jie Shen, Xuan Deng, Chao Zhang, Ming Guan

## Abstract

Diffuse large B-cell lymphoma (DLBCL) is the predominant type of central nervous system lymphoma (CNSL) including primary CNSL and secondary CNSL. Diffuse large B cells in cerebrospinal fluid (CSF-DLBCs) have offered great promise for the diagnostics and therapeutics of CNSL leptomeningeal involvement. To explore the distinct phenotypic states of CSF-DLBCs, we analyzed the transcriptomes of 902 CSF-DLBCs from six CNSL-DLBCL patients using single-cell RNA sequencing technology. We defined CSF-DLBCs based on abundant expression of B-cell markers, as well as the enrichment of cell proliferation and energy metabolism pathways. CSF-DLBCs within individual patients exhibited monoclonality with similar variable region of light chains (VL) expression. It is noteworthy that we observed some CSF-DLBCs have double classes of VL (lambda and kappa) transcripts. We identified substantial heterogeneity in CSF-DLBCs, and found significantly greater among-patient heterogeneity compared to among-cell heterogeneity within a given patient. The transcriptional heterogeneity across CSF-DLBCs is manifested in cell cycle state and cancer-testis antigens expression. Our results will provide insight into the mechanism research and new diagnostic direction of CNSL-DLBCL leptomeningeal involvement.

## Introduction

Primary central nervous system lymphoma (PCNSL) is a rare and aggressive extranodal non-Hodgkin lymphoma (NHL), accounting for up to 1% of NHL and about 3% of all primary brain tumors^1^. PCNSL is confined to the brain, eyes, spinal cord, or leptomeninges without systemic involvement and regarded as an “immune-privileged (IP)” lymphoma^2^. The majority of PCNSL cases (>95%) are diffuse large B□cell lymphoma (DLBCL) with expression of B-cell markers^3^. In contrast to PCNSL-DLBCL, systemic DLBCL lymphoma at diagnosis or relapse involved both within and outside the CNS is defined as secondary CNS lymphoma DLBCL (SCNSL-DLBCL)^4^. Due to the poor central nervous system penetration of drugs and the prolonged overall survival of patients, the incidence of CNS-DLBCL has been increasing in recent decades.

For patients with suspected CNS lymphoma, a histopathologic diagnosis by stereotactic brain biopsy is the gold standard^5^. However, brain biopsy is an invasive method with a risk of complications, and decreased sensitivity of biopsies as a result of the administration of corticosteroids can delay the initiation of systemic therapy^6^. Considerable recent improvements in neuroimaging techniques have provided the requisite sensitivity for diagnosis of CNSL and are able to define the site and extent of the disease, but neuroimaging findings are not specific^7^. In recent years, multimodal investigations of cerebrospinal fluid (CSF) have greatly facilitated CNSL diagnosis. Positive cytopathological examination of the CSF is still regarded as the “gold standard” for a definitive diagnosis of CNSL leptomeningeal involvement^8^. In addition, the advent of immunophenotypic, molecular genetic mutations and interleukins further expand the diagnostic value of CSF^9,10^. CSF obtained through lumbar puncture has the advantage of minimal risk and can be sampled multiple times, which is beneficial to monitor the progress of CNSL. The treatment of CNSL has evolved from the use of whole-brain radiotherapy alone to multimodality regimens that include chemotherapy with high-dose methotrexate, monoclonal antibodies, and autologous stem cell transplantation^11,12^. The development of new technologies and approaches has improved the diagnosis and therapy of CNSL, but the overall prognosis for PCNSL remains relatively poor with a mean survival period of less than 5 years^13^.

Until now, a comprehensive understanding of CNSL-DLBCL mechanisms is still lacking. To make breakthroughs in tackling the clinical challenge of CNSL-DLBCL, we intended to analyze the transcriptome characteristics of diffuse large B cells (DLBCs) in CSF (CSF-DLBCs) from CNSL-DLBCL patients, which is of great significance for the discovery of new diagnostic biomarkers and development of novel targeted therapeutic approaches. Traditional RNA sequencing of tumor cells in bulk is confounded by the presence of other cells and unable to capture gene expression heterogeneity among tumor cells. In order to avoid the disadvantages of RNA sequencing in bulk, we obtained the transcriptional profiles of CSF-DLBCs from CNSL-DLBCL patients by Smart-seq2 single-cell RNA sequencing (scRNA-seq). In this study, we investigated systematic and comprehensive characterization of more than nine hundred CSF-DLBCs at the single-cell transcriptome level for the first time.

### Profiles of individual cells in CSF

In this study, we enrolled patient CSF samples from six CNSL-DLBCL patients (P1-P6), 3 normal CSF samples (N1-N3), and some blood T/B cells from one healthy volunteer (supplemental Table 1). Normal CSF samples (N1-N3) were collected from patients who were screened for potential brain infections and diagnosed as normal. Normal CSF samples were sorted by Calcein Blue AM positivity to enrich live cells. Patient CSF samples were diagnosed through cytopathology (supplemental Figure 1A). CSF-DLBCs are CD45 and CD19 positive, and much larger in morphology than normal cells with greater forward scatter height FSC-H in FACS (supplemental Figure 1B). In addition to CSF cells, blood T/B cells were sorted and sequenced to help define the CSF cells’ composition. Individual live target cells were sorted for scRNA-seq library preparation using the SMART-seq2 approach. The libraries were sequenced on an Illumina HiSeqX system to achieve on average 300,000 reads per cell. The data were analyzed using Seurat 3.1.1. We performed scRNA-seq on 2,727 target cells and kept 1,527 cells with high quality transcriptome data for subsequent analysis (Table 1).

**Table 1.** Summary of cell type identity in scRNA-seq results of cancer patient CSF samples. PCNSL: primary central nervous system lymphoma; SCNSL: secondary central nervous system lymphoma; DLBCL: diffuse large B-cell lymphoma.

| Patient ID | Diagnostics | Number of sequenced cells | Number of QC filtered cells | T cells | B cells | Monocytes | Diffuse large B cells |
| --- | --- | --- | --- | --- | --- | --- | --- |
| P1 | SCNSL-DLBCL | 519 | 306 | 0 | 0 | 1 | 305 |
| P2 | PCNSL-DLBCL | 36 | 31 | 0 | 0 | 9 | 22 |
| P3-1 | PCNSL-DLBCL | 348 | 85 | 43 | 4 | 35 | 3 |
| P3-2 |  | 384 | 220 | 4 | 0 | 1 | 213 |
| P4 | PCNSL-DLBCL | 480 | 167 | 25 | 6 | 0 | 132 |
| P5 | PCNSL-DLBCL | 48 | 40 | 1 | 3 | 2 | 34 |
| P6 | SCNSL-DLBCL | 336 | 195 | 0 | 0 | 2 | 193 |
| N1 | Control | 288 | 175 | 144 |  | 28 |  |
| N2 | Control | 240 | 109 | 103 |  | 3 |  |
| N3 | Control | 96 | 37 | 24 |  | 11 |  |
| Blood-T cells | Normal | 168 | 82 | 80 |  |  |  |
| Blood-B cells | Normal | 168 | 80 |  | 79 |  |  |
| **Total** |  | **3,111** | **1,527** | **424** | **92** | **92** | **902** |

**Figure 1.**
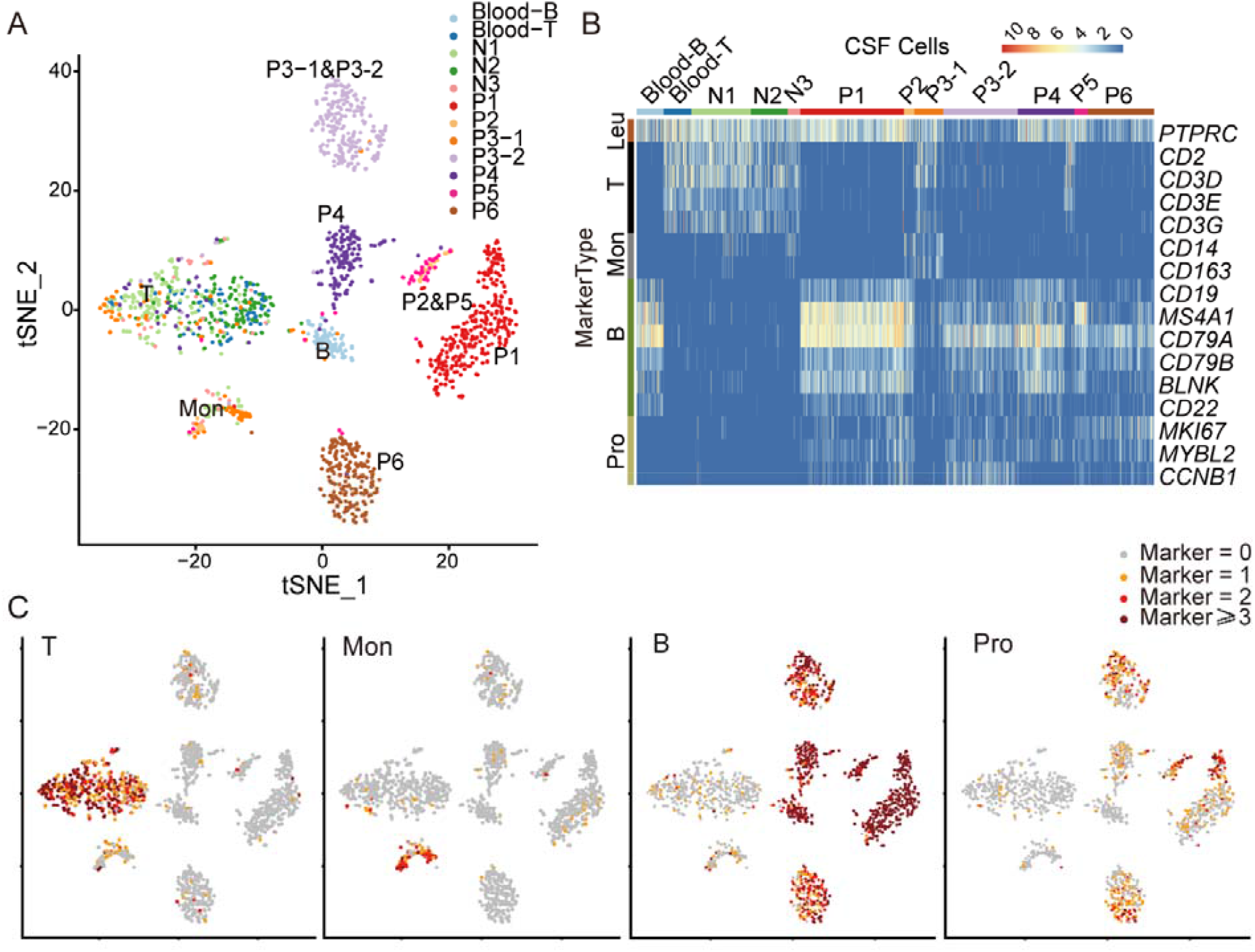
Clustering and analysis of single-cell expression data of CSF samples. (A) 2D representation of samples correlations by t-SNE dimensionality reduction including six CNSL-DLBCL CSF samples (P), three normal CSF samples (N), blood T and B cells. (B) Heat map showing expression of selected gene panels in different samples. (C) Feature plots demonstrating the expression of selected gene panels on the t-SNE plot (Figure 1A). Scaled expression levels are depicted by the number of expressing markers. No marker expressing, gray; one marker expressing, orange; two markers expressing number, red; more than two markers expressing number, dark red. Leukocyte (Leu) marker gene: *PTPRC*; T-cell (T) marker genes: *CD2, CD3D/E/G*; Monocytes (Mon) marker genes: *CD14, CD68, CD163*; B-cell (B) marker genes: *CD19, MS4A1, CD79A, CD79B, BLNK, CD22*; Proliferation (Pro) associated genes: *MKI67, MYBL2, CCDB1*.

CSF cells were clustered by t-distributed stochastic neighbor embedding (t-SNE) ^14^. On the basis of their preferentially or distinctively expressed marker genes (Figure 1A-C; supplemental Figure 2), three clusters of nonmalignant cells were annotated as T cells (424 cells), B cells (92 cells), or Monocytes (92 cells; Table 1). Interestingly, blood-T and CSF-T cells, or blood-B and CSF-B cells are in the same cluster (Figure 1A), indicating that normal lymphocytes have similar expression profiles in different microenvironments. To confirm this conclusion, normal lymphocytes were clustered individually and still separated into three clusters (supplemental Figure 3).

**Figure 2.**
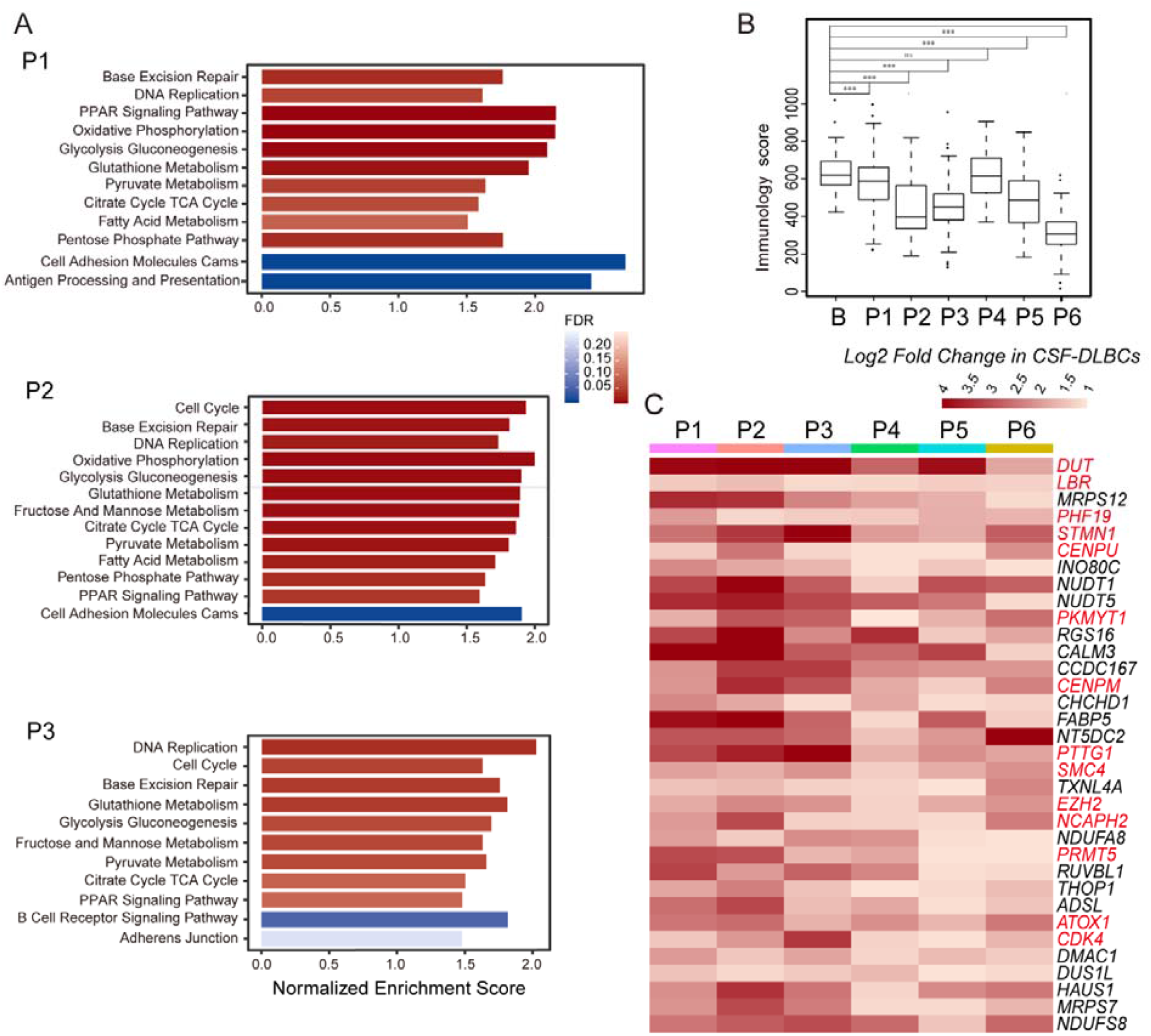
Characteristics of CSF-DLBCs using single-cell transcriptome analysis. (A) GSEA analysis showing significantly upregulated (red gradient) or downregulated (blue gradient) KEGG pathways in CSF-DLBCs of patient P1, P2 and P3 compared to normal B cells (*P* < 0.05). (B) The immune signature of cells quantified by the ImmuneScore computed from the ESTIMATE algorithm, showing the significant difference between the B-cell group and the patient CSF-DLBCs group (***: *P* < 0.001, Wilcoxon Rank-Sum test). (C) Heat map showing the 34 selected genes (supplemental Table 3) upregulated in six patient CSF-DLBCs compared to normal B cells (adjusted *P*-value < 0.05; log2(fold-change) ≥1) and expressed in fewer than 5% normal B cells.

**Figure 3.**
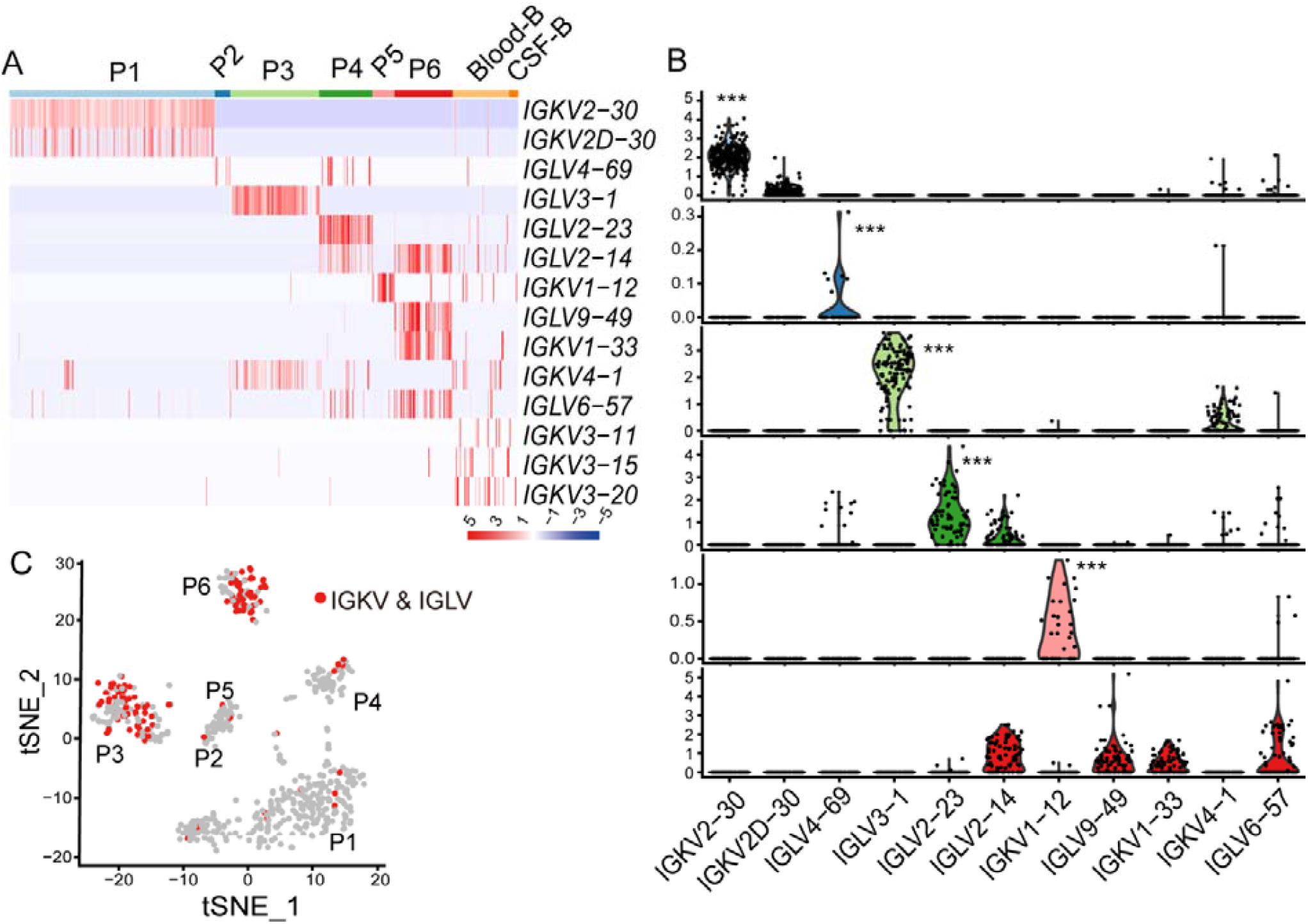
Characterization of immunoglobulin light chain variable molecules in CSF-DLBCs. (A) Heatmap displaying the distribution of expression of immunoglobulin light chain variable molecules in CSF-DLBCs (IGKV, immunoglobulin kappa variable; IGLV, immunoglobulin lambda variable). (B) Violin plots displaying the distribution of expression of immunoglobulin light chain variable molecules in different patient CSF-DLBCs (***: *P* < 0.001, Wilcoxon Rank-Sum test). (C) t-SNE plot of 624 CSF-DLBCs with more than 1000 covered genes (supplemental Figure 5A). Cells are colored red expressing both IGKV and IGLV.

The majority of patient CSF cells strongly clustered according to the patient of origin with the exception of normal leukocytes (Figure 1A). Patient (P) P3-1 and P3-2 samples were collected from the same patient within a two-month time interval. The proportion of DLBCs in the P3-1 CSF sample was 2% by cytopathological analysis, whereas in the P3-2 CSF sample it was 60%, indicating tumor progression during the two months. The P3-1 sample had three cells (the rest are leukocytes) clustered with the P3-2 sample despite that they had undergone independent cell sorting, library construction and sequencing (Figure 1A). We did not observe significant heterogeneity in mapping quality and gene coverage across patient samples. The clustering pattern is not driven by technical variability and batch effect.

### Transcriptome signatures of diffuse large B cells in CSF

At the molecular level, we have defined 902 CSF-DLBCs (P1, 305; P2, 22; P3, 216; P4, 132; P5, 34; P6, 132; Table 1) with transcriptome signatures for B-cell markers and proliferation markers (Figure 1B-C; supplemental Figure 2), especially *MKI67*, a classical proliferation marker commonly used in clinical immunohistochemistry.

We performed gene set enrichment analysis (GSEA) to further determine the functional enrichment in CSF-DLBCs compared to normal B cells. We discovered that cell proliferation category was significantly enriched in CSF-DLBCs from six patients (*P*<0.05; Figure 2A; supplemental Figure 4). The cell proliferation category consists of cell cycle, DNA replication and repair, further confirming that CSF-DLBCs are in an actively proliferative state. The metabolism pathway category was also enriched in CSF-DLBCs, significantly from P1, P2 and P3 (*P*<0.05; Figure 2A; supplemental Figure 4). The metabolism pathway category mainly contains pentose phosphate, glycolysis gluconeogenesis, pyruvate metabolism, citrate cycle TCA cycle, glutathione metabolism, fatty acid metabolism and PPAR (proliferator-activated receptor) signaling pathway. These energy metabolism pathways are critical for tumor growth and the energy demand in the brain. In addition, the enriched energy metabolism pathways of patient P4, P5 and P6 CSF-DLBCs were fewer than that of other patients (Figure 2A; supplemental Figure 4).

**Figure 4.**
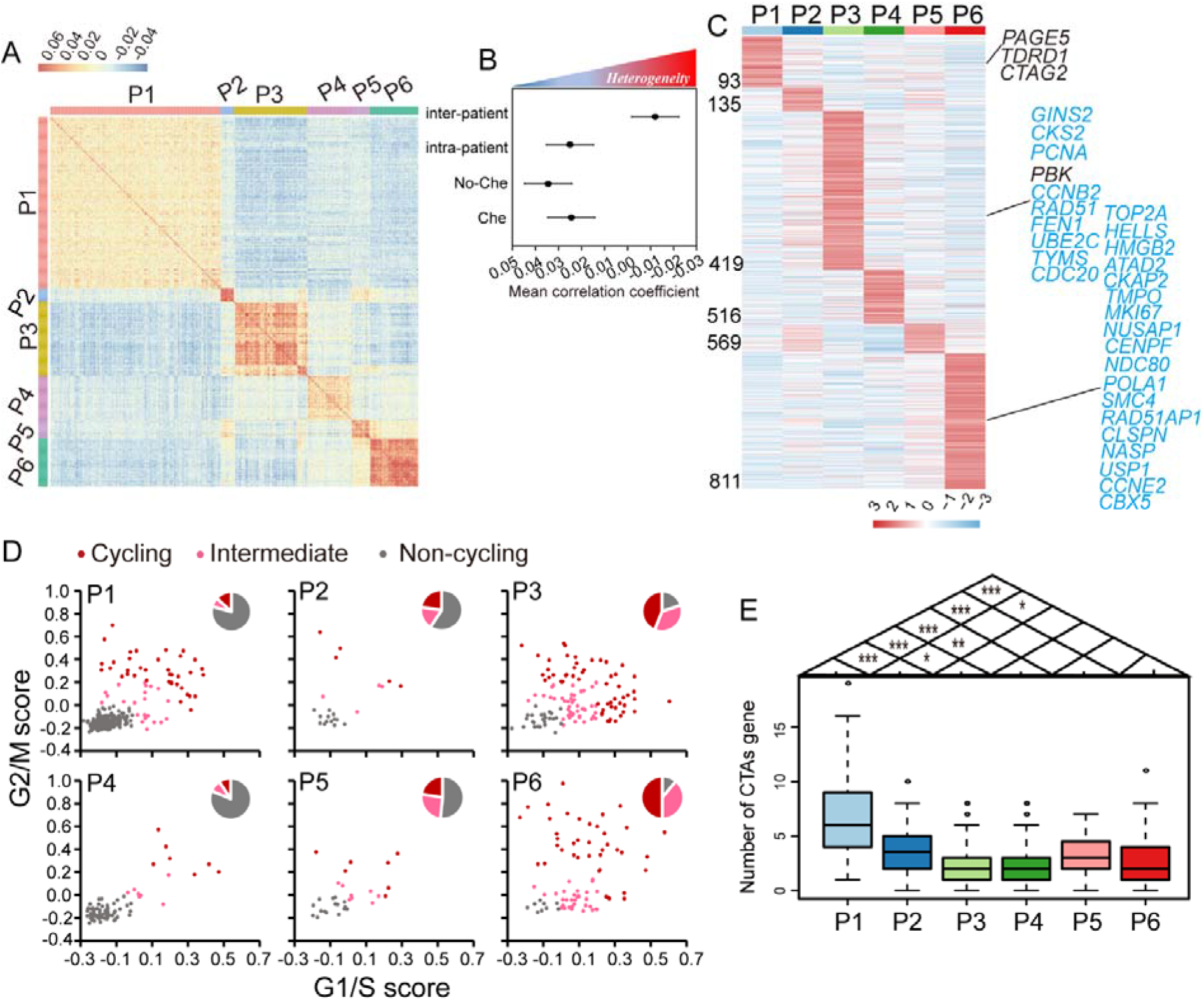
The heterogeneity of CSF-DLBCs among different patients and within individual patient. (A) The pairwise correlations between the expression profiles of CSF-DLBCs from six CNSL-DLBCL patients. (B) heterogeneity analysis showing the mean correlation coefficients for CSF-DLBCs within individual CNSL-DLBCL patient (intra-patient), among CNSL-DLBCL patient (inter-patient), and for CSF-DLBCs within individual CNSL-DLBCL patient with chemotherapy (P1, P3, P6; Che) or without chemotherapy (P2, P4, P5; No-Che). (C) Heatmap of differentially expressed genes (adjusted *P*-value < 0.05, fold change >1.5) that are exclusively or preferentially expressed in one individual CNSL-DLBCL patient. The names of selected genes are labeled. Gene names marked in black (blue) are cell cycle related genes (cancer-testis antigens; CTAs). (D) Estimation of the cell cycle state of every CSF-DLBC (circles) on the basis of relative expression of G1/S (x axis) and G2/M (y axis) gene sets in different CNSL-DLBCL patient. Cells are colored by their inferred cell cycle states: cycling cells (score>0.2), red; intermediate (0<score≤0.2), pink; and noncycling cells (score≤0), gray. (E) Boxplot of the number of expressed CTAs (y-axis) in CSF-DLBCs from six CNSL-DLBCL patient (x-axis).

The observed down-regulation of the antigen processing and presentation pathway and the B cell receptor signaling pathway in CSF-DLBCs (Figure 2A; supplemental Figure 4) suggested their decreased capacity as immune cells. To further delineate immune characteristics of CSF-DLBCs, the ImmuneScore was computed based on ESTIMATE R package^15^. The result showed normal B cells have higher ImmuneScore than CSF-DLBCs, except those from P4 (Figure 2B).

To address the fundamental differences in the expression program of the pure CSF-DLBCs population, we employed the DESeq2 method to assess whether a gene is differentially expressed between normal B cells and CSF-DLBCs within each patient. Overall, 3582 genes were identified as differential expression genes (DEGs) in at least 1 sample, and 173 DEGs were identified in all patients (adjusted *P*-value < 0.05; | log2(fold-change)| ≥1). In order to determine the genes that were specifically expressed in CSF-DLBCs, we selected out 34 genes upregulated in six patients and detected in fewer than 5% of normal B cells (Figure 2C; supplemental Table 2). Among these, *EZH2* (Enhancer of zeste homolog 2), whose overexpression and mutation has been identified in many cancers including non-Hodgkin lymphomas, acts to increase *H3K27* (histone H3 at lysine 27) methylation and promote tumor progression^16,17^. Additionally, the upregulated expression of *PRMT5* (protein arginine methyltransferase 5) is required for the survival and proliferation of DLBCL^18^. In addition, *DUT*^*19*^, *LBR*^*20*^, *PHF19*^*21*^, *STMN1*^*22*^, *CENPU*^*23*^, *CENPM*^*24*^, *PKMYT1*^*25*^, *NCAPH2*^*26*^, *PTTG1*^*27*^, *SMC4*^*28*^, *ATOX1*^*29*^ and *CDK4*^*30*^ have also been reported to play important roles in cell cycle and proliferation. These 34 genes deserve further study due to their expression specificity in CSF-DLBCs.

### The analysis of variable region of light chain (VL) in CSF-DLBCs

When analyzing the differences of CSF-DLBCs between six CNSL-DLBCL patients, we further removed cells that had fewer than 1000 genes, and 624 CSF-DLBCs were retained (P1, 289; P2, 22; P3, 125; P4, 75; P5, 31; P6, 82; supplemental Figure 5). The immunoglobulin light chain restriction (LCR) indicates monoclonality of the proliferating mature B cells. CSF-DLBCs of the same patient had similar variable region of light chain (VL) expression (Figure 3A). The VL molecule mainly expressed in P1 is IGKV2-30 (kappa VL, IGKV), in P2 is IGLV4-69 (lambda VL, IGLV), in P3 is IGLV3-1, in P4 is IGLV2-23, and in P5 is IGKV1-12 (Figure 3A-B). Additionally, other VL molecules were detected in CSF-DLBCs with lower expression (Figure 3A-B). P6 was atypical in that four VL molecules were expressed similarly in CSF-DLBCs (lambda VL, IGLV2-14, IGLV9-49, IGLV6-57; kappa VL, IGKV1-33; Figure 3A-B).

As we know, DLBCL exhibits allelic exclusion in which only a single class of light chain is expressed (either lambda λ or kappa κ). However, there have been increasing numbers of reports that a double class of light-chain gene rearrangements can occur in B-cell malignant neoplasms^31^. P6 is such a sample with 60.98% (50/82) of CSF-DLBCs expressing κ and λ light chains (Figure 3C). P3 also had 58/125 CSF-DLBCs expressing dual κ/λ light chains (Figure 3C), though the transcription level of IGKV4-1 was lower than that of IGLV3-1. The remainder of the patients had few CSF-DLBCs with dual κ/λ light chains expression (P1, 9/289; P2, 1/22; P4, 7/75; P5, 2/31; Figure 3C).

For VL expression of normal B cells, normal B cells were polyclonal and only 6/92 cells had dual light-chain transcriptions (Figure 3A), which is consistent with previous reports that 0.2%-3.4% of normal maturing B cells have dual light-chain expression ^32,33^.

### Gene expression heterogeneities of CSF-DLBCs

The analysis of cell-to-cell correlation showed significant heterogeneity between CSF-DLBCs within an individual patient in spite of the monoclonality of VL (correlation coefficients ranging from −0.043 to 0.693; Figure 4A). The correlation between CSF-DLBCs within an individual patient is much higher than that among different patients (mean correlation coefficient −0.012 vs. 0.025, *P*-value < 2.2e-16, Wilcoxon Rank-Sum test; Figure 4A-B).

CSF-DLBCs from patients who had undergone chemotherapy (P1, P3, P6) showed considerably greater intercellular heterogeneity than those from patients who had not received chemotherapy (P2, P4, P5), which is consistent with the view that chemotherapy promotes the progression of tumor cells (mean coefficient 0.0244 vs. 0.0345, *P*-value < 2.2e-16, Wilcoxon Rank-Sum test; Figure 4B). As shown in Figure 1A, CSF-DLBCs cluster of patient with chemotherapy is closer to B-cell cluster than that of patient without chemotherapy.

### Cell cycle heterogeneity of CSF-DLBCs

A total of 811 differentially expressed genes were identified in individual CSF-DLBCs (*P*-value < 0.05, fold change >1.5; Figure 4C). From the differential gene expression list, we can see many cell cycle related genes upregulated in P3 and P6 (Figure 4C). To characterize this different proliferation state of CSF-DLBCs, we used gene signatures to denote G1/S or G2/M phases^34^. Cell cycle phase-specific signatures were highly expressed in a subset of CSF-DLBCs, distinguishing cycling cells from noncycling cells (supplemental Figure 6A). These signatures revealed variability in the fraction of cycling cells across six patients; the proportion of CSF-DLBCs in the cycling state is higher in P3 (44%) and P6 (50%) than in the other patients (Figure 4D). In addition, compared to non-cycling cells, the cell cycle genes *TOP2A, CCNB2, CDC20* and *SMC4* were greatly upregulated in cycling cells regardless of the tumor proliferation state of the patient (supplemental Figure 6B-C). These genes are candidates for proliferation markers with MKI67 in the diagnosis of CSF-DLBCs.

### Cancer-testis antigens (CTAs) heterogeneity of CSF-DLBCs

The differential gene expression list among patients also included cancer-testis antigens (CTAs). CTAs have particular characteristics of high immunogenicity with restricted expression in normal tissues, and provide unprecedented opportunities for cancer diagnosis and immunotherapy^35^. Several studies have evaluated the expression of CTAs in non-Hodgkin’s lymphoma to date^36,37^, but little is known about the expression of CTAs in CSF-DLBCs. We examined the expression of 276 selected CTAs (http://www.cta.lncc.br/modelo.php) in CSF-DLBCs and discovered substantial inter-tumor heterogeneity and intra-tumor heterogeneity of CTAs (Figure 4C; supplemental Figure 7). The number of expressed CTAs in CSF-DLBCs is different among patients, and patient P1 exhibited expression of the most CTAs (Figure 4E). In addition, the expression of *PAGE5, TDRD1, CTAG2, MAEL, CT45A1, PAGE2B* and *MAGEA9B* are almost restricted to P1 CSF-DLBCs (supplemental Figure 7). *ATAD2* (29%, 181/624) and *MPHOSPH10* (29.17%, 182/624) are ubiquitously and highly expressed in CSF-DLBCs among all patients, and have the potential to serve as immunotherapy targets (supplemental Figure 7).

## Discussion

In this study, we analyzed the transcriptomes of CSF-DLBCs at the single cell level from 6 CNSL-DLBCL patients. CSF-DLBCs mainly clustered according to patient of origin, whereas P2 and P5 CSF-DLBCs were in the same cluster, which was characterized by high expression of many ribosomal genes (data not shown). Although P2 and P5 CSF-DLBCs clustered together, they had different variable region of light chain molecules (Figure 3A-B). Immunoglobulin light chain restriction (LCR) is a feature of DLBCs; individual patient DLBCs showed monoclonality of VL and three CSF-DLBCs in the P3-1 sample expressed IGLV3-1, which was the same as CSF-DLBCs in the P3-2 sample. Therefore, we analyzed the transcriptomes of CSF-DLBCs based on individual patients.

ScRNA-Seq enabled us to directly compare the transcriptomes of entirely pure normal B cells and CSF-DLBCs. Compared to normal B cells, gene expression in CSF-DLBCs was enriched in the cell proliferation and energy metabolism pathways, which are critical for tumor growth and energy demand in the CSF-DLBCs. As reported, consensus cluster classification has grouped DLBCLs into the B cell receptor (BCR)/proliferation cluster (BCR-DLBCL), the OxPhos cluster (OxPhos-DLBCL), and the host response (HR) cluster with a brisk host inflammatory infiltrate^38^. Compared to BCR-DLBCL, OxPhos-DLBCL have enhanced oxidative phosphorylation, tricarboxylic acid cycle (TCA), fatty acid oxidation program (FAO), PPAR signaling pathway, pyruvate metabolism, glucose-derived metabolites and glutathione synthesis, but do not display active/functional BCR signaling^39^. Based on the active metabolism characteristics of CSF-DLBCs, CNS-DLBCL might be grouped into the OxPhos-DLBCL cluster with down-regulated B cell receptor signaling pathway, especially patients P1, P2 and P3 (Figure 2A; supplemental Figure 4). As is well known, adhesive cell-cell and cell-matrix interactions generally play important roles in tumor metastases and drug resistance^40^. Whether the generally down-regulated cell adhesion molecules (CAMs) pathway in CSF-DLBCs affects CNS-DLBCL metastases and drug resistance deserves further study.

### Heterogeneities of CSF-DLBCs

We identified the transcriptional heterogeneity of CSF-DLBCs in cell cycle state and the expression of cancer-testis antigens. In addition, heterogeneities were also observed in other aspects. First, the enriched pathways of energy metabolism in CSF-DLBCs are different among patients to some extent. Second, HLA-II (human leukocyte antigen class II) molecules (*HLA-DRB5, HLA-DQA1, HLA-DQB1, HLA-DMB, HLA-DRA*) were highly expressed in P3 and P4, but not in other patients (supplemental Figure 8). The frequent loss of HLA-II in CNS-DLBCL is mainly due to homozygous deletions in the HLA region, which affects the onset and modulation of immune response for lack of activated *CD4*^+^ T lymphocytes^41^. Last, DLBCL can be divided into germinal center B-cell-like (GCB) and non-GCB, and up to 96% of PCNSL-DLBCL are classified as non-GCB type^10^. Based on the expression of *MME* (*CD10*), *BCL6* and *IRF4* (*MUM1*) molecules (Hans algorithm), the individual CSF-DLBCs have both GCB and non-GCB subtypes (supplemental Figure 9). Our classification only depended on RNA level not immunohistochemistry, but showed heterogeneity of CSF-DLBCs from one aspect. In addition, there was no obvious transcriptional difference between PCNSL-DLBCL and SCNSL-DLBCL, both of them have extensive heterogeneity.

Until now, the intricate machinery of CSF-DLBCs immune evasion has been still unknown, thus analyzing the immune cell characteristics in pathological CSF samples would be informative to understand a systems-level view of the CNS-DLBCL tumor microenvironment. A recent single-cell RNA-seq study of follicular lymphoma tumor microenvironment revealed the coexpression of immune checkpoints in regulatory T (Treg) cells, offering a better understanding of immune regulation^42^. Another scRNA-seq study made reported that the infiltrating T cells of lung adenocarcinoma have great heterogeneity and patients with a low ratio of “pre-exhausted” to exhausted T cells have worse prognosis^43^. While our study solely focused on CSF-DLBCs, the characteristics of immune cells in CNSL-DLBCL patient CSF are worthy of future research.

To date, there are scant CSF scRNA-seq studies in the literature. One identified rare CSF immune cell subsets of patients with HIV infection were able to perpetuate neuronal injury^44^. The other study revealed that multiple sclerosis increases CSF cell type diversity including cytotoxic phenotype T helper cells, and follicular cells expanded B lineage cells in CSF^45^. Our study is the first to analyze the transcriptome characteristics of CSF-DLBCs at single-cell resolution. Despite analyzing a small number of patients, our study revealed novel biological insights of the intricate machinery responsible for CNS-DLBCL progression, and helped to develop new approaches for CNS-DLBCL diagnosis and immunotherapy. However, a larger validation cohort is required in the future.

## Methods

### Patients information

All human sample materials used in this study were collected with informed consent. The proposed studies were approved by the Ethics Committee of Huashan Hospital, Fudan University. Clinical information of patients is listed in supplemental Table 1.

### Single-cell preparation

Antibody (CD45, CD3, CD19, BD Biosciences) and labeling dye for live cells (Calcein Blue AM, Life Technologies, CA) were used per manufacturer recommendations. Live cells (Calcein Blue AM+) in normal CSF samples (N: N1-N3) and live tumor cells (Calcein Blue AM+, CD45+, CD19+) in patient CSF samples (P: P1-P6) were collected for sequencing (supplemental Table 1). Blood-T cells (Calcein Blue AM+, CD45+, CD3+) and blood-B cells (Calcein Blue AM+, CD45+, CD19+) were also sorted for sequencing (supplemental Table 1). Target cells were sorted into pre-prepared 96-well plates by FACS (fluorescence-activated cell sorting). Single-cell lysates were sealed, vortexed, centrifuged, immediately placed on dry ice and transferred for storage at −80°C.

### SMART-seq2 library construction

Library for isolated single cell was generated by SMART-Seq2 method ^46^ with the following modifications: RNA was reverse transcribed with Maxima H Minus Reverse Transcriptase (Thermo Fisher Scientific, MA), and whole transcriptome was amplified using KAPA HiFi Hot Start Ready Mix (KAPA Biosystems, MA). cDNA library was purified using Agencourt XP DNA beads (Beckman Coulter, CA) and quantified with a high sensitivity dsDNA Quant Kit (Life Technologies, CA). It is worth mentioning that full length cDNA libraries were tagmented, and then only 3’ end sequence (500-1000bp) was amplified and enriched for sequencing on an Illumina HiSeqX machine, which is different from the traditional SMART-Seq2 method of full tagmented-libraries sequence.

### Generation of gene expression matrix

Sequenced reads were mapped to hg38 using the STAR (version 2.7; https://github.com/alexdobin/STAR) with the default parameters. These uniquely mapped reads in the genome were used, and reads aligned to more than one locus were discarded. The expression level of gene was quantified by the number of counts.

Then, in the gene expression matrix from 12 samples, genes expressed (counts > 0) in less than 10 cells were filtered out. Cells were removed according to the following criteria: (1) cells had fewer than 500 genes; (2) cells had over 15% mitochondrial-gene counts. A filtered gene expression matrix including 1,527 cells and 11,289 genes were used in the following analysis (R package Seurat 3.1.1; https://cran.r-project.org/web/packages/Seurat/index.html). In addition, when analyzing the differences of CSF-DLBCs between six CNSL-DLBCL patients, we further removed cells that had fewer than 1000 genes and 624 CSF-DLBCs were retained.

### Population identification

After filtration, a merged expression matrix of 12 samples was used for cell clustering by the Seurat package (version 3.1.1), adapting the typical pipeline. In brief, gene expression was normalized by the “NormalizeData” function. Highly variable genes were calculated with the “FindVariableGenes” method with the default parameters. Significant principal components were used for downstream graph-based, semi-unsupervised clustering into distinct populations (FindClusters function in R) and t-SNE dimensionality reduction was used to project these populations in two dimensions. To identify marker genes, the clusters were compared pairwise for differential gene expression using the Wilcoxon rank-sum test for single-cell gene expression (FindAllMarkers function, min.pct = 0.1, logFC.threshold = 0.25). Subsequently, cell clusters were annotated manually, based on known markers.

### Analysis of differential expression and gene enrichment

DESeq2 (R package DESeq2 v3.9; https://bioconductor.org/packages/release/bioc/html/DESeq2.html) was also used to detect DEGs (differential expression genes) between target samples^47^. GSEA (gene set enrichment analysis) was used for functional enrichment analysis of KEGG pathways^48^.

### Cell cycle analysis

Cell cycle assignment was performed in R using the CellCycleScoring function included package “Seurat” (https://cran.r-project.org/web/packages/Seurat/index.html). We identified cells that had either S.Score or G2M.Score > 0.2 as cycling cells; cells that had either 0 <S.Score or G2M.Score ≤0.2 as intermediate cells; and the other cells as non-cycling cells.

## Data Availability

The single-cell RNA sequencing data will be submitted to NCBI Sequence Read Archive.

## Acknowledgements

This work is supported by the National Key Research and Development Program of China (Grant No. 2017YFA0103902), the National Natural Science Foundation of China (Grant No. 31771283), the Fundamental Research Funds for the Central Universities (Grant No. 22120190210), the Innovation Group Project of Shanghai Municipal Health Commission (2019CXJQ03). M.G. is supported by the Program for Shanghai Municipal Leading Talent (2015).

## References

1. Villano JL, Koshy M, Shaikh H, Dolecek TA, McCarthy BJ. Age, gender, and racial differences in incidence and survival in primary CNS lymphoma. Br J Cancer. 2011;105(9):1414–1418.

2. Han CH, Batchelor TT. Diagnosis and management of primary central nervous system lymphoma. Cancer. 2017;123(22):4314–4324.

3. Giannini C, Dogan A, Salomao DR. CNS lymphoma: a practical diagnostic approach. J Neuropathol Exp Neurol. 2014;73(6):478–494.

4. Baraniskin A, Chomiak M, Ahle G, et al. MicroRNA-30c as a novel diagnostic biomarker for primary and secondary B-cell lymphoma of the CNS. J Neurooncol. 2018;137(3):463–468.

5. Khatab S, Spliet W, Woerdeman PA. Frameless image-guided stereotactic brain biopsies: emphasis on diagnostic yield. Acta Neurochir (Wien). 2014;156(8):1441–1450.

6. Onder E, Arikok AT, Onder S, et al. Corticosteroid pre-treated primary CNS lymphoma: a detailed analysis of stereotactic biopsy findings and consideration of interobserver variability. Int J Clin Exp Pathol. 2015;8(7):7798–7808.

7. Nabavizadeh SA, Vossough A, Hajmomenian M, Assadsangabi R, Mohan S. Neuroimaging in Central Nervous System Lymphoma. Hematol Oncol Clin North Am. 2016;30(4):799–821.

8. Baraniskin A, Schroers R. Modern cerebrospinal fluid analyses for the diagnosis of diffuse large B-cell lymphoma of the CNS. CNS Oncol. 2014;3(1):77–85.

9. Sasagawa Y, Akai T, Tachibana O, Iizuka H. Diagnostic value of interleukin-10 in cerebrospinal fluid for diffuse large B-cell lymphoma of the central nervous system. J Neurooncol. 2015;121(1):177–183.

10. Hiemcke-Jiwa LS, Leguit RJ, Snijders TJ, et al. Molecular analysis in liquid biopsies for diagnostics of primary central nervous system lymphoma: Review of literature and future opportunities. Crit Rev Oncol Hematol. 2018;127:56–65.

11. Chukwueke UN, Nayak L. Central Nervous System Lymphoma. Hematol Oncol Clin North Am. 2019;33(4):597–611.

12. Nayak L, Batchelor TT. Recent advances in treatment of primary central nervous system lymphoma. Curr Treat Options Oncol. 2013;14(4):539–552.

13. Camilleri-Broet S, Criniere E, Broet P, et al. A uniform activated B-cell-like immunophenotype might explain the poor prognosis of primary central nervous system lymphomas: analysis of 83 cases. Blood. 2006;107(1):190–196.

14. van der Maaten L, Hinton G. Visualizing Data using t-SNE. Journal of Machine Learning Research. 2008;9:2579–2605.

15. Yoshihara K, Shahmoradgoli M, Martinez E, et al. Inferring tumour purity and stromal and immune cell admixture from expression data. Nat Commun. 2013;4:2612.

16. Chase A, Cross NC. Aberrations of EZH2 in cancer. Clin Cancer Res. 2011;17(9):2613–2618.

17. Morin RD, Mendez-Lago M, Mungall AJ, et al. Frequent mutation of histone-modifying genes in non-Hodgkin lymphoma. Nature. 2011;476(7360):298–303.

18. Lu X, Fernando TM, Lossos C, et al. PRMT5 interacts with the BCL6 oncoprotein and is required for germinal center formation and lymphoma cell survival. Blood. 2018;132(19):2026–2039.

19. Vilpo JA, Autio-Harmainen H. Uracil-DNA glycosylase and deoxyuridine triphosphatase: studies of activity and subcellular location in human normal and malignant lymphocytes. Scand J Clin Lab Invest. 1983;43(7):583–590.

20. Arai R, En A, Takauji Y, et al. Lamin B receptor (LBR) is involved in the induction of cellular senescence in human cells. Mech Ageing Dev. 2019;178:25–32.

21. Ning F, Wang C, Niu S, Xu H, Xia K, Wang N. Transcription factor Phf19 positively regulates germinal center reactions that underlies its role in rheumatoid arthritis. Am J Transl Res. 2018;10(1):200–211.

22. Baik SY, Yun HS, Lee HJ, et al. Identification of stathmin 1 expression induced by Epstein-Barr virus in human B lymphocytes. Cell Prolif. 2007;40(2):268–281.

23. Zhang Q, Li YD, Zhang SX, Shi YY. Centromere protein U promotes cell proliferation, migration and invasion involving Wnt/beta-catenin signaling pathway in non-small cell lung cancer. Eur Rev Med Pharmacol Sci. 2018;22(22):7768–7777.

24. Xiao Y, Najeeb RM, Ma D, Yang K, Zhong Q, Liu Q. Upregulation of CENPM promotes hepatocarcinogenesis through mutiple mechanisms. J Exp Clin Cancer Res. 2019;38(1):458.

25. Schmidt M, Rohe A, Platzer C, Najjar A, Erdmann F, Sippl W. Regulation of G2/M Transition by Inhibition of WEE1 and PKMYT1 Kinases. Molecules. 2017;22(12).

26. Wallace HA, Rana V, Nguyen HQ, Bosco G. Condensin II subunit NCAPH2 associates with shelterin protein TRF1 and is required for telomere stability. J Cell Physiol. 2019;234(11):20755–20768.

27. Huang JL, Cao SW, Ou QS, et al. The long non-coding RNA PTTG3P promotes cell growth and metastasis via up-regulating PTTG1 and activating PI3K/AKT signaling in hepatocellular carcinoma. Mol Cancer. 2018;17(1):93.

28. Steffensen S, Coelho PA, Cobbe N, et al. A role for Drosophila SMC4 in the resolution of sister chromatids in mitosis. Curr Biol. 2001;11(5):295–307.

29. Matson Dzebo M, Blockhuys S, Valenzuela S, Celauro E, Esbjorner EK, Wittung-Stafshede P. Copper Chaperone Atox1 Interacts with Cell Cycle Proteins. Comput Struct Biotechnol J. 2018;16:443–449.

30. Hosooka T, Ogawa W. A novel role for the cell cycle regulatory complex cyclin D1-CDK4 in gluconeogenesis. J Diabetes Investig. 2016;7(1):27–28.

31. Xu D. Dual surface immunoglobulin light-chain expression in B-cell lymphoproliferative disorders. Arch Pathol Lab Med. 2006;130(6):853–856.

32. Diaw L, Siwarski D, DuBois W, Jones G, Huppi K. Double producers of kappa and lambda define a subset of B cells in mouse plasmacytomas. Mol Immunol. 2000;37(12-13):775–781.

33. Giachino C, Padovan E, Lanzavecchia A. kappa+lambda+ dual receptor B cells are present in the human peripheral repertoire. J Exp Med. 1995;181(3):1245–1250.

34. Tirosh I, Izar B, Prakadan SM, et al. Dissecting the multicellular ecosystem of metastatic melanoma by single-cell RNA-seq. Science. 2016;352(6282):189–196.

35. Salmaninejad A, Zamani MR, Pourvahedi M, Golchehre Z, Hosseini Bereshneh A, Rezaei N. Cancer/Testis Antigens: Expression, Regulation, Tumor Invasion, and Use in Immunotherapy of Cancers. Immunol Invest. 2016;45(7):619–640.

36. Hudolin T, Kastelan Z, Ilic I, et al. Immunohistochemical analysis of the expression of MAGE-A and NY-ESO-1 cancer/testis antigens in diffuse large B-cell testicular lymphoma. J Transl Med. 2013;11:123.

37. Inaoka RJ, Jungbluth AA, Gnjatic S, et al. Cancer/testis antigens expression and autologous serological response in a set of Brazilian non-Hodgkin’s lymphoma patients. Cancer Immunol Immunother. 2012;61(12):2207–2214.

38. Monti S, Savage KJ, Kutok JL, et al. Molecular profiling of diffuse large B-cell lymphoma identifies robust subtypes including one characterized by host inflammatory response. Blood. 2005;105(5):1851–1861.

39. Caro P, Kishan AU, Norberg E, et al. Metabolic signatures uncover distinct targets in molecular subsets of diffuse large B cell lymphoma. Cancer Cell. 2012;22(4):547–560.

40. Wu Y, Xu X, Miao X, et al. Sam68 regulates cell proliferation and cell adhesion-mediated drug resistance (CAM-DR) via the AKT pathway in non-Hodgkin’s lymphoma. Cell Prolif. 2015;48(6):682–690.

41. Jordanova ES, Philippo K, Giphart MJ, Schuuring E, Kluin PM. Mutations in the HLA class II genes leading to loss of expression of HLA-DR and HLA-DQ in diffuse large B-cell lymphoma. Immunogenetics. 2003;55(4):203–209.

42. Andor N, Simonds EF, Czerwinski DK, et al. Single-cell RNA-Seq of follicular lymphoma reveals malignant B-cell types and coexpression of T-cell immune checkpoints. Blood. 2019;133(10):1119–1129.

43. Guo X, Zhang Y, Zheng L, et al. Global characterization of T cells in non-small-cell lung cancer by single-cell sequencing. Nat Med. 2018;24(7):978–985.

44. Farhadian SF, Mehta SS, Zografou C, et al. Single-cell RNA sequencing reveals microglia-like cells in cerebrospinal fluid during virologically suppressed HIV. JCI Insight. 2018;3(18).

45. Schafflick D, Xu CA, Hartlehnert M, et al. Integrated single cell analysis of blood and cerebrospinal fluid leukocytes in multiple sclerosis. Nat Commun. 2020;11(1):247.

46. Picelli S, Bjorklund AK, Faridani OR, Sagasser S, Winberg G, Sandberg R. Smart-seq2 for sensitive full-length transcriptome profiling in single cells. Nat Methods. 2013;10(11):1096–1098.

47. Love MI, Huber W, Anders S. Moderated estimation of fold change and dispersion for RNA-seq data with DESeq2. Genome Biology. 2014;15(12).

48. Subramanian A, Tamayo P, Mootha VK, et al. Gene set enrichment analysis: a knowledge-based approach for interpreting genome-wide expression profiles. Proc Natl Acad Sci U S A. 2005;102(43):15545–15550.

